# Left Atrial Stiffness Trajectories Identify Distinct Prognostic Phenotypes in Heart Failure with Reduced Ejection Fraction

**DOI:** 10.64898/2026.05.20.26353741

**Authors:** Jonghee Sun, Jiesuck Park, Nan Young Bae, Jaehyun Lim, Soongu Kwak, MinJung Bak, Hong-Mi Choi, Jun-Bean Park, Yeonyee E. Yoon, Seung-Pyo Lee, Yong-Jin Kim, Goo-Yeong Cho, Hyung-Kwan Kim, In-Chang Hwang

**Affiliations:** Department of Cardiology, Cardiovascular Center, Seoul National University Bundang Hospital, Republic of Korea; Department of Internal Medicine, Seoul National University College of Medicine, Republic of Korea; Healthcare System Gangnam Center, Seoul National University Hospital, Republic of Korea; Cardiovascular Center and Department of Internal Medicine, Seoul National University Hospital, Republic of Korea

**Keywords:** angiotensin receptor–neprilysin inhibitor, heart failure with reduced ejection fraction, left atrial stiffness index, left atrial reservoir strain, reverse remodeling, atrial fibrillation

## Abstract

**Background:** Treatment response in heart failure with reduced ejection fraction (HFrEF) is assessed predominantly through left ventricular (LV) functional recovery, while longitudinal changes in left atrial (LA) hemodynamic burden remain underexplored. The LA stiffness index (LASI), derived from E/e′ and LA reservoir strain, integrates LV filling pressure and LA compliance.

**Objectives:** We investigated longitudinal trajectories of LASI and their prognostic implications in HFrEF treated with angiotensin receptor–neprilysin inhibitor (ARNI)-based therapy.

**Methods:** From the multicenter STRATS-HF-ARNI registry, 1,039 patients with HFrEF who underwent serial echocardiography at baseline and one-year follow-up were classified into four LASI trajectory patterns dichotomized at the cohort median (1.22): persistently compliant (Group A, 46.8%), reverse remodeling (B, 28.5%), progressive stiffening (C, 3.2%), and persistently stiff (D, 21.6%).

**Results:** On multivariable Cox regression, Group D was independently associated with elevated risks of all-cause mortality (adjusted hazard ratio [aHR] 2.68, 95% CI 1.57–4.59), cardiovascular mortality (aHR 4.36, 1.97–9.64), and HF hospitalization (aHR 3.83, 2.22–6.60), whereas Group B showed outcomes comparable to Group A. One-year LASI progression independently predicted all three outcomes. LASI elevation at one year predicted adverse outcomes even among patients with recovered LV function, and LASI trajectory classification provided incremental prognostic discrimination beyond conventional diastolic and strain parameters. Among sinus-rhythm patients (n=786), Group C exhibited the highest risk of new-onset atrial fibrillation.

**Conclusions:** In HFrEF treated with ARNI-based therapy, LASI trajectories identify distinct prognostic phenotypes. Persistent LA stiffness confers adverse outcomes independent of LV recovery, and serial LASI assessment may enhance risk stratification beyond LV-centric metrics.

**CONDENSED ABSTRACT:** In 1,039 patients with HFrEF treated with ARNI-based therapy, four longitudinal left atrial stiffness index (LASI) trajectories were identified using serial echocardiography. Persistent LA stiffness was independently associated with elevated all-cause mortality, cardiovascular mortality, and HF hospitalization. The prognostic value of one-year LASI persisted even among patients with recovered LV function, and LASI trajectory provided incremental prognostic discrimination beyond conventional diastolic and strain parameters. Progressive stiffening of an initially compliant left atrium identified the highest risk of new-onset atrial fibrillation. Serial assessment of LA stiffness may enhance risk stratification beyond LV-centric metrics in HFrEF.

## INTRODUCTION

Assessment of treatment response in heart failure (HF) with reduced ejection fraction (HFrEF) has traditionally relied on left ventricular (LV)-centric parameters, such as left ventricular ejection fraction (LVEF). Contemporary HF therapy, including angiotensin receptor–neprilysin inhibitor (ARNI), has improved clinical outcomes and promoted favorable reverse remodeling in HFrEF.^1–3^ However, improvement in LVEF does not necessarily indicate complete resolution of HF pathobiology, as patients continue to experience worsening symptoms, abnormal biomarker profiles, and adverse events despite apparent LV recovery.^4,5^ Therefore, additional markers that capture residual hemodynamic burden beyond LV recovery may improve risk stratification.

The left atrium (LA) is chronically exposed to elevated LV filling pressure and reflects the cumulative hemodynamic burden of HF. Conventional echocardiographic indices, such as E/e′ and LA volume index (LAVI), are widely used to assess LV filling pressure and chronic LA remodeling, but LA enlargement alone may not fully reflect LA mechanical dysfunction.^6^ LA reservoir strain (LASr) provides functional information on LA capacity and compliance, and impaired LASr has been associated with adverse outcomes in HFrEF.^7–9^ In this context, the LA stiffness index (LASI), calculated as E/e′ divided by LASr, integrates LV filling pressure and LA reservoir function into a single index reflecting LA compliance.^10,11^ Elevated LASI is associated with adverse clinical outcomes in HF and may help identify elevated LV filling pressure in acute decompensation.^11–13^

Despite growing interest in LA mechanical assessment, the clinical relevance of LASI has been studied mainly in HF patients with preserved LVEF (HFpEF), while its prognostic value in HFrEF patients receiving ARNI-based therapy remains insufficiently characterized. Moreover, little is known about serial changes in LASI during treatment, including whether LA stiffness improves in parallel with LV recovery and whether persistent or progressive LA stiffness identifies residual risk despite apparent LV functional improvement.

Therefore, using a multicenter HFrEF registry, we investigated longitudinal trajectories of LASI in patients treated with ARNI-based therapy. Specifically, we (1) classified patients according to baseline and one-year LASI patterns and characterized their clinical and hemodynamic features across these patterns, and (2) evaluated the associations between LASI patterns and clinical outcomes, including mortality, HF hospitalization, and new-onset atrial fibrillation (AF).

## METHODS

### Study Population

This study used data originated from the STrain for Risk Assessment and Therapeutic Strategies in patients with Heart Failure treated with Angiotensin Receptor–Neprilysin Inhibitor (STRATS-HF-ARNI) registry, a multicenter observational cohort. The registry comprised 2,757 consecutive patients with a confirmed diagnosis of heart failure with reduced ejection fraction (HFrEF) who received ARNI-based HF therapy at two tertiary referral centers in the Republic of Korea (Seoul National University Hospital and Seoul National University Bundang Hospital) from 2017 through 2022. The registry was registered with the Clinical Research Information Service under the Ministry of Health and Welfare of the Republic of Korea (registration number: KCT0008098); the registry methodology and overall patient characteristics have been reported in prior publications.^14,15^ The current analysis was focused to patients who had completed serial transthoracic echocardiographic assessments at both the index visit and the one-year follow-up. Patients were excluded for the following reasons: no baseline echocardiographic assessment (n=336), no one-year follow-up echocardiographic assessment (n=1,218), or insufficient echocardiographic data (n=164). The resulting analytic cohort comprised 1,039 patients (**Figure 1**). Ethical approval was obtained from the Institutional Review Boards at each participating site (Seoul National University Hospital: J-2212-034-1383; Seoul National University Bundang Hospital: B-2005-615-108), and the requirement for individual informed consent was waived given the retrospective observational nature of the study.

**Figure 1.**
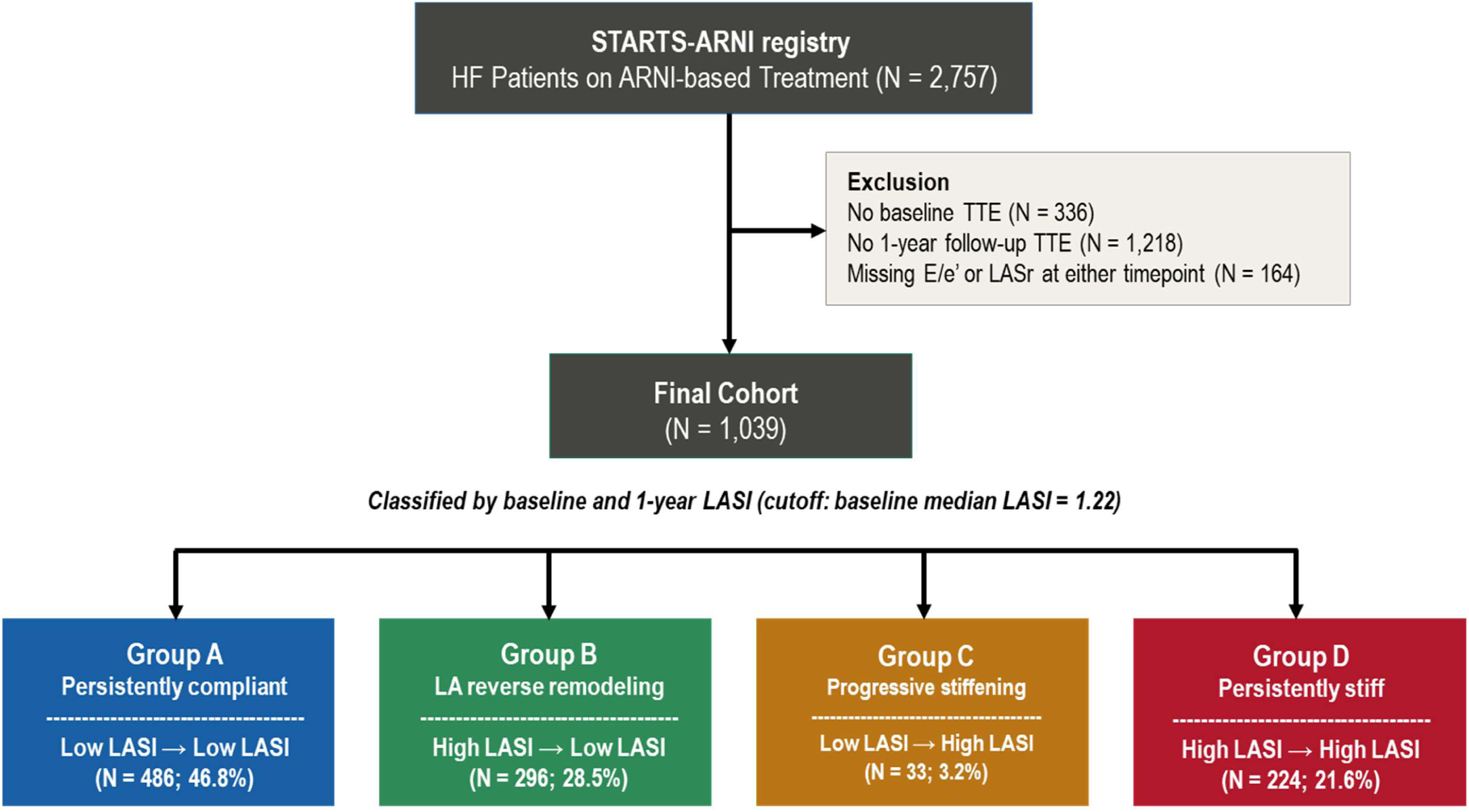
Study Flow. Of 2,757 consecutive patients with HFrEF on ARNI-based treatment in the STRATS-HF-ARNI registry, 1,039 patients with available E/e′ and LASr measurements at both baseline and one-year follow-up were included in the final analysis. Patients were classified into four mutually exclusive LASI trajectory patterns according to baseline and one-year LASI values, each dichotomized at the cohort median: Group A, persistently compliant; Group B, LA reverse remodeling; Group C, progressive stiffening; and Group D, persistently stiff. ARNI, angiotensin receptor–neprilysin inhibitor; E/e′, ratio of early transmitral flow velocity to early diastolic mitral annular velocity; HFrEF, heart failure with reduced ejection fraction; LA, left atrium; LASI, left atrial stiffness index; LASr, left atrial reservoir strain; TTE, transthoracic echocardiography.

### Echocardiographic Evaluation

Serial echocardiographic examinations were conducted as part of standard clinical practice by credentialed sonographers under the supervision of cardiologists with specialized expertise in cardiac imaging, in accordance with contemporary guideline recommendations.^6^ Dedicated TTE assessments were obtained at baseline and at the one-year follow-up visit.

Biplane LVEF was derived using the modified Simpson’s method, integrating planimetric measurements from the apical four-chamber and two-chamber views. LAVI was determined from the maximal left atrial volume referenced to body surface area, measured at end-systole from the same apical views. The E/e′ ratio was obtained by dividing the peak early transmitral inflow velocity by the septal early diastolic mitral annular velocity, acquired by pulsed-wave Doppler imaging. LASr was derived from two-dimensional speckle-tracking analysis performed with dedicated software (TomTec Image Arena 4.6; TomTec Imaging Systems, Munich, Germany), and was defined as the peak positive longitudinal deformation of the LA myocardium, measured from the apical four-chamber and two-chamber views.^16^ Higher LASr values reflect greater LA reservoir function and more preserved LA compliance.

### Definition of LA Stiffness Index and Cutoff Determination

LASI was calculated as E/e′ divided by the absolute value of LASr, integrating LV filling pressure and LA compliance into a single composite index.^10,11,17^ A higher LASI reflects the combination of elevated LV filling pressure and impaired LA reservoir function, representing a greater stiffness-related hemodynamic burden on the LA. Although previous studies have proposed LASI cutoffs ranging from 0.26 to 0.76,^11–13^ these values varied substantially according to study setting and HF phenotype, with most evidence derived from HFpEF populations, and have not been established in serial assessments of HFrEF patients under ARNI-based therapy. To explore the clinical relevance of longitudinal LASI changes in the present cohort, the baseline median LASI value was adopted as the cutoff for pattern classification. Accordingly, LA status was classified as compliant when baseline LASI was below the cohort median, and as stiff when baseline LASI was at or above the cohort median.

### Classification of LASI Trajectory Patterns

Four mutually exclusive LASI trajectory groups were defined by cross-classifying patients according to their baseline and one-year LASI values relative to the cohort-derived median cutoff: Group A (compliant at both baseline and one year; persistently compliant), Group B (stiff at baseline but compliant LA features at one year; LA reverse remodeling), Group C (compliant at baseline but stiff at one year; progressive stiffening), and Group D (stiff at both baseline and one year; persistently stiff). Group A served as the reference category in all comparative analyses.

### Clinical Outcomes

The primary outcome was all-cause mortality. Secondary outcomes included cardiovascular mortality and HF hospitalization. Additionally, the risk of new-onset AF was assessed in the subset of patients in sinus rhythm without a history of AF at baseline. New-onset AF was defined as the first documentation of AF on routine clinical electrocardiography or Holter monitoring. Clinical outcomes were ascertained through dedicated review of the electronic medical records. The one-year follow-up echocardiography served as the landmark index date for all outcome analyses.

### Statistical Analysis

Clinical data are presented as median with interquartile range (IQR) for continuous variables and as frequency with percentage for categorical variables. Differences in baseline characteristics across the four groups were assessed using the Kruskal-Wallis test for continuous variables and the chi-square test for categorical variables.

Serial changes between baseline and one-year follow-up were assessed within each group using the Wilcoxon signed-rank test. The parameters analyzed included LASI, its components (E/e′ and LASr), and hemodynamic variables (LVEF, LAVI, PASP, and log-transformed N-terminal pro–B-type natriuretic peptide [NT-proBNP]). Between-group differences in the magnitude of change were evaluated using the Kruskal-Wallis test. The association between the change in LVEF and the change in LASI was examined using Spearman’s rank correlation coefficient.

Cumulative incidence curves were constructed using the Kaplan-Meier estimator and compared between groups by the log-rank test. The independent association between LASI trajectory group and each clinical endpoint was examined through multivariable Cox proportional hazards regression, with Group A designated as the reference. Covariates included in all models were age, sex, hypertension, diabetes mellitus, chronic kidney disease, AF, baseline LVEF, baseline LAVI, and medication use (beta-blocker, mineralocorticoid receptor antagonist, and sodium-glucose cotransporter-2 inhibitor). Effect estimates are reported as adjusted hazard ratios (aHR) with 95% confidence intervals (95% CI). In a complementary analysis, the one-year change in LASI (ΔLASI) was treated as a continuous predictor to characterize the dose-response relationship between the magnitude of LA stiffness change and subsequent outcome risk. To evaluate the prognostic value of LASI independent of LV functional recovery, two complementary analyses were additionally performed. First, patients were cross-classified by LV recovery status at one year, defined as ΔLVEF ≥10% from baseline, and by one-year LASI status (below vs. at or above the cohort median). Second, patients were stratified by one-year LVEF category (<40%, ≥40 to <50%, and ≥50%) and by one-year LASI status. In both analyses, the association between one-year LASI status and clinical outcomes was assessed using the same multivariable model.

For the analysis of new-onset AF, patients in sinus rhythm at baseline without a history of AF were included. The association between LASI trajectory pattern and new-onset AF was evaluated using Kaplan-Meier estimation and the same multivariable Cox regression model. To further examine how the magnitude of LASI improvement related to AF risk beyond categorical group comparisons, the joint association of ΔLASI and one-year LASI with new-onset AF was additionally assessed using a multivariable Cox regression model with restricted cubic splines for both variables, and the resulting hazard ratios were visualized as a heatmap. The reference point was set at no change in LASI (ΔLASI = 0) and the cohort median of baseline LASI.

All statistical analyses were performed using Python (version 3.12.3; Python Software Foundation). Two-sided p values <0.05 were considered statistically significant.

## RESULTS

### Study Population and Baseline Characteristics

A total of 1,039 patients with HFrEF receiving ARNI-based therapy were included in the final analysis. The distributions of baseline E/e′, LASr, and LASI in the overall cohort are shown in **Figure S1**. Baseline LASI exhibited a median of 1.22 (IQR, 0.72–2.42), and this cohort-specific median value was adopted as the cutoff for trajectory pattern classification. Patients were classified into Group A (persistently compliant, n=486, 46.8%), Group B (LA reverse remodeling, n=296, 28.5%), Group C (progressive stiffening, n=33, 3.2%), and Group D (persistently stiff, n=224, 21.6%) (**Figure 1**).

Baseline characteristics across the four groups are summarized in **Table 1**. Patients in Groups C (74.0 [IQR 64.0–77.6] years) and D (71.7 [62.2–77.3] years) were older than those in Groups A (63.9 [54.6–72.9] years) and B (66.2 [56.2–75.1] years, p<0.001), without significant differences in sex distribution. Hypertension and diabetes mellitus were most prevalent in Groups C and D (hypertension, 57.6% and 37.5%; diabetes mellitus, 48.5% and 34.8%, respectively). Prevalence of chronic kidney disease and AF were highest in Group D (36.6% and 43.3%, respectively), followed by Group B (25.3% and 28.0%). NT-proBNP levels were lowest in Group A (627 [258–1650] pg/mL) and markedly elevated in Groups B and D (2,469 [1129–5422] and 3,387 [1441–7539] pg/mL, respectively).

**Table 1.** Baseline Characteristics by LASI Pattern.

|  | Group A<br>Persistently compliant<br>(Low → Low LASI) | Group B<br>LA reverse remodeling<br>(High → Low LASI) | Group C<br>Progressive stiffening<br>(Low → High LASI) | Group D<br>Persistently stiff<br>(High → High LASI) | p |
| --- | --- | --- | --- | --- | --- |
| N | 486 | 296 | 33 | 224 |  |
| Age, year | 63.9 (54.6–72.9) | 66.2 (56.2–75.1) | 74.0 (64.0–77.6) | 71.7 (62.2–77.3) | <0.001 |
| Male | 350 (71.9) | 202 (68.2) | 23 (69.7) | 143 (63.8) | 0.191 |
| BMI, kg/m<sup>2</sup> | 24.7 (22.3–27.1) | 24.1 (22.0–26.8) | 23.9 (21.8–24.6) | 24.3 (22.3–26.2) | 0.241 |
| Comorbidities |  |  |  |  |  |
| Hypertension | 158 (32.4) | 98 (33.1) | 19 (57.6) | 84 (37.5) | 0.020 |
| Diabetes mellitus | 114 (23.4) | 80 (27.0) | 16 (48.5) | 78 (34.8) | <0.001 |
| Chronic kidney disease | 84 (17.2) | 75 (25.3) | 8 (24.2) | 82 (36.6) | <0.001 |
| Atrial fibrillation | 69 (14.2) | 83 (28.0) | 4 (12.1) | 97 (43.3) | <0.001 |
| Medications at baseline |  |  |  |  |  |
| Beta-blocker | 456 (93.6) | 272 (91.9) | 27 (81.8) | 189 (84.4) | <0.001 |
| MRA | 226 (46.4) | 166 (56.1) | 22 (66.7) | 111 (49.6) | 0.016 |
| SGLT2 inhibitor | 80 (16.4) | 63 (21.3) | 7 (21.2) | 35 (15.6) | 0.258 |
| Laboratory |  |  |  |  |  |
| eGFR, mL/min/1.73m<sup>2</sup> | 79.6 (60.4–94.1) | 68.1 (52.7–87.8) | 68.7 (52.9–87.5) | 63.6 (42.9–81.7) | <0.001 |
| NT-proBNP, pg/mL | 627 (258–1650) | 2469 (1129–5422) | 1619 (870–3577) | 3387 (1441–7539) | <0.001 |
| Baseline Echocardiography |  |  |  |  |  |
| LV ejection fraction, % | 33.1 (28.1–37.0) | 26.8 (22.0–32.5) | 32.0 (27.4–36.4) | 29.8 (24.5–34.0) | <0.001 |
| LV end-diastolic dimension, mm | 59.0 (54.0–63.0) | 61.0 (55.0–66.0) | 59.0 (55.0–65.0) | 59.0 (54.6–64.0) | 0.005 |
| LA volume index, mL/m<sup>2</sup> | 44.8 (34.8–56.7) | 61.6 (50.3–77.5) | 49.5 (40.1–69.9) | 76.0 (56.5–91.7) | <0.001 |
| E/e' ratio | 11.7 (9.1–14.6) | 20.9 (16.0–27.8) | 15.0 (10.8–16.5) | 24.0 (18.0–30.7) | <0.001 |
| LA reservoir strain, % | 17.6 (14.5–21.9) | 9.1 (7.0–12.0) | 16.1 (14.2–17.6) | 8.3 (6.2–10.8) | <0.001 |
| PASP, mmHg | 28.0 (24.4–33.3) | 41.0 (30.0–51.0) | 30.0 (27.0–36.7) | 43.0 (33.0–53.4) | <0.001 |
| LV global longitudinal strain, % | -11.1 (-13.1–-9.0) | -8.3 (-10.5–-6.1) | -9.2 (-12.0–-8.0) | -8.5 (-10.8–-6.7) | <0.001 |
| Baseline LASI | 0.71 (0.50–0.93) | 2.17 (1.58–3.33) | 0.85 (0.72–1.06) | 2.93 (1.86–4.38) | <0.001 |

Data are presented as median [IQR] for continuous variables and n (%) for categorical variables. p values from Kruskal-Wallis test (continuous) or chi-square test (categorical).
Abbreviations: AF, atrial fibrillation; BMI, body mass index; eGFR, estimated glomerular filtration rate; LA, left atrium; LASI, LA stiffness index; LV, left ventricle; MRA, mineralocorticoid receptor antagonist; NT-proBNP, N-terminal pro-B-type natriuretic peptide; PASP, pulmonary artery systolic pressure; SGLT2i, sodium-glucose cotransporter-2 inhibitor.

Baseline echocardiographic parameters differed significantly across groups. Groups B and D exhibited more advanced hemodynamic derangement than Groups A and C, with elevated E/e′ (20.9 [16.0–27.8] and 24.0 [18.0–30.7]), enlarged LAVI (61.6 [50.3–77.5] and 76.0 [56.5–91.7] mL/m²), reduced LASr (9.1% [7.0–12.0] and 8.3% [6.2–10.8]), and elevated PASP (41.0 [30.0–51.0] and 43.0 [33.0–53.4] mmHg). LVGLS also showed the greatest impairment in Groups B and D (−8.3% [−10.5 – −6.1] and −8.5% [−10.8 – −6.7]) compared with Groups A and C (−11.1% [−13.1 – −9.0] and −9.2% [−12.0 – −8.0]). By design, baseline LASI differed markedly across groups, with median values of 0.71, 2.17, 0.85, and 2.93 in Groups A, B, C, and D, respectively.

### Serial Changes in LASI and Its Components

Serial trajectories of LASI from baseline to one-year follow-up differed markedly across the four groups (**Figure 2A**). In Group B, LASI decreased substantially from a median of 2.17 (IQR, 1.58–3.33) to 0.62 (IQR, 0.42–0.85). Conversely, Group C showed a marked increase in LASI from a baseline median of 0.85 (IQR, 0.72–1.06) to 1.59 (IQR, 1.42–2.00) at one year. Group D remained persistently elevated without meaningful reduction. Group A maintained consistently low values below the cutoff throughout follow-up. Examination of the two LASI components (E/e’ and LASr) revealed distinct recovery patterns (**Figure 2B**). In Group B, both components improved concordantly, with E/e′ decreasing by a median of 9.6 and LASr increasing by 9.2. In Group C, the changes occurred in the opposite direction, with E/e′ increasing by 5.2 and LASr decreasing by 6.3. Groups A and D exhibited smaller directional changes consistent with their stable trajectory classifications. The relationship between one-year changes in LV and LA function demonstrated that ΔLVEF and ΔLASI were inversely correlated (Spearman r=−0.39, p<0.001; **Figure 2C**), indicating that greater LV functional recovery was associated with greater LASI reduction. However, the magnitude of correlation indicated substantial residual variance, suggesting that LVEF recovery alone does not fully capture LA functional recovery.

**Figure 2.**
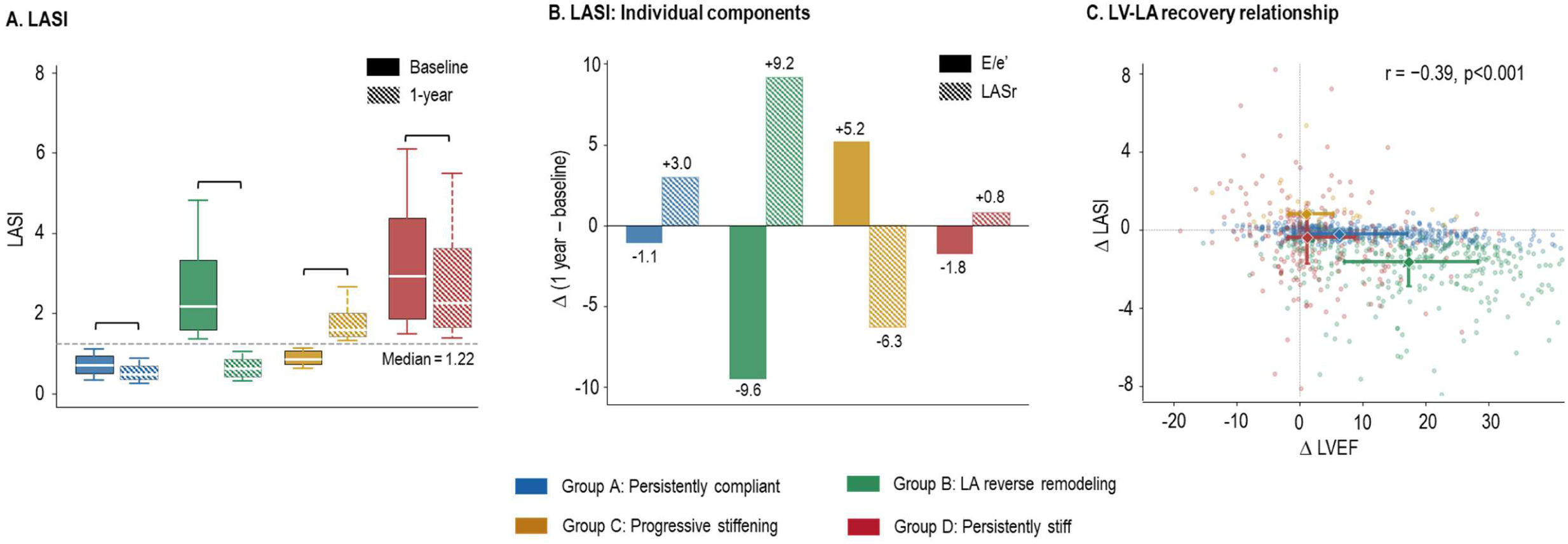
Temporal Trends in LASI Patterns. (A) Serial trajectories of LASI from baseline to one-year follow-up by LASI trajectory group, presented as median with interquartile range. The dashed line indicates the cohort median (LASI = 1.22) used as the cutoff for group classification. (B) Between-group differences in ΔLASI and its components (ΔE/e′ and ΔLASr). (C) Scatter plot of ΔLVEF against ΔLASI, with the Spearman correlation coefficient shown. E/e′, ratio of early transmitral flow velocity to early diastolic mitral annular velocity; LASI, left atrial stiffness index; LASr, left atrial reservoir strain; LVEF, left ventricular ejection fraction.

Regarding the baseline determinants of LASI improvement at 1-year, multivariable logistic regression identified both favorable and unfavorable predictors (**Table 2**). Older age (OR per 10-year increment: 0.88, 95% CI 0.78–0.99, p=0.031) and diabetes mellitus (OR: 0.56, 95% CI 0.34–0.92, p=0.023) independently predicted failure of LASI normalization, while higher baseline E/e′ (OR per 1-unit: 1.02, 95% CI 1.00–1.03, p=0.010) and lower baseline LASr (OR per 1% decrease: 1.09, 95% CI 1.04–1.13, p<0.001) each independently favored LASI recovery. Baseline LVEF, LAVI, and β-blocker use were associated with LASI improvement on univariable analysis but lost significance after multivariable adjustment.

**Table 2.**
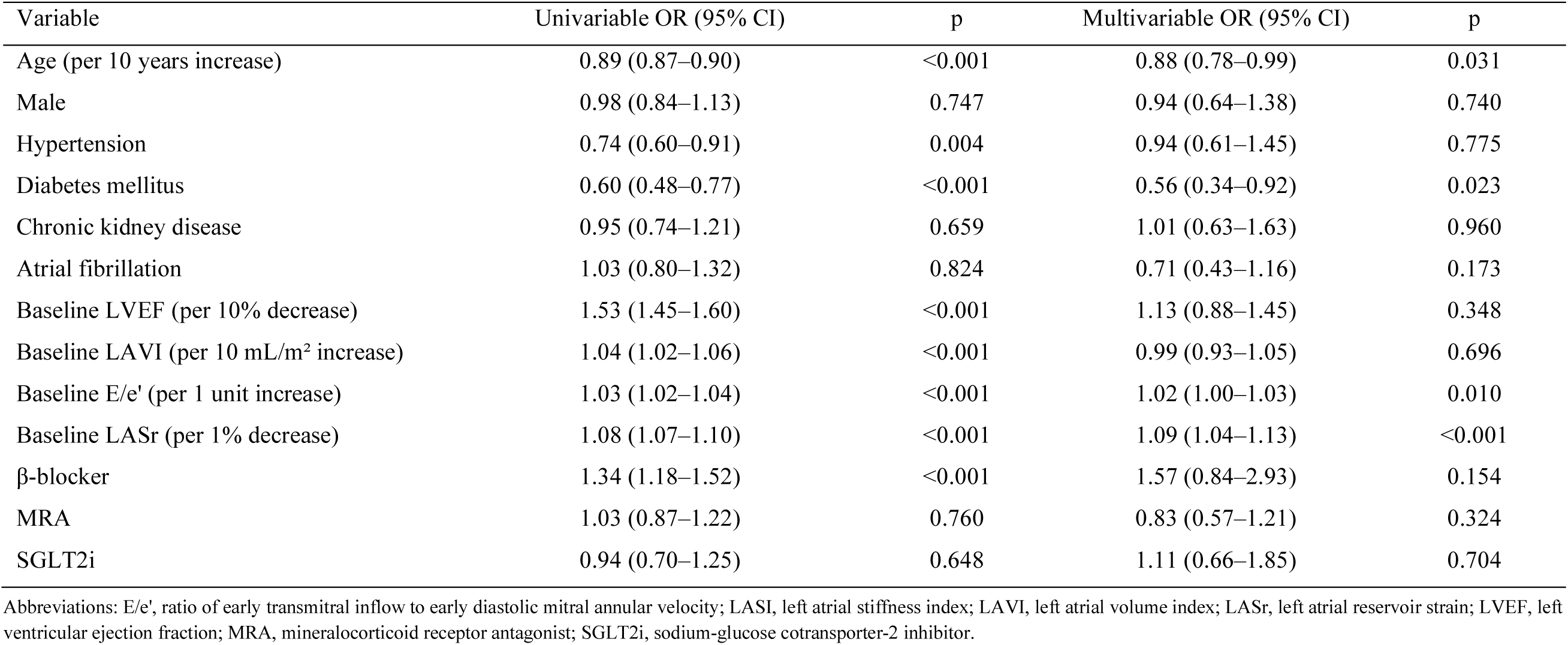
Predictors of LASI Improvement at 1-year Period.

### Hemodynamic Trajectories by LASI Pattern

Longitudinal changes in hemodynamic parameters across the four groups are shown in **Table 3**. LVEF improved from baseline to one year in all groups, with the greatest magnitude of recovery observed in Group B. LAVI showed substantial reduction in Group B, whereas Group C exhibited an increasing trend and Group D showed persistent LA enlargement without meaningful improvement. E/e′ decreased markedly in Group B, increased significantly in Group C, and remained elevated in Group D. PASP and log-transformed NT-proBNP showed analogous patterns, with favorable improvement in Group B and persistent elevation in Group D. Between-group differences were statistically significant for all parameters (p<0.001 for all).

**Table 3.** Serial Hemodynamic and Echocardiographic Changes by LASI Trajectory Group.

| Variable | Timepoint | Group A<br>Persistently compliant | Group B<br>LA reverse remodeling | Group C<br>Progressive stiffening | Group D<br>Persistently stiff | p value* |
| --- | --- | --- | --- | --- | --- | --- |
| LVEF (%) | Baseline | 33.1 (28.1–37.0) | 26.8 (22.0–32.5) | 32.0 (27.4–36.4) | 29.8 (24.5–34.0) | <0.001 |
|  | 1-year | 41.0 (35.0–50.0) | 45.7 (36.0–54.7) | 35.0 (29.1–40.0) | 32.0 (26.1–38.0) | <0.001 |
|  | Δ (1yr–baseline) | 6.2 (1.1–17.0) | 17.3 (7.0–28.2) | 1.0 (-1.8–5.2) | 1.1 (-2.0–8.6) | <0.001 |
|  | Within-group p† | <0.001 | <0.001 | 0.118 | <0.001 |  |
| LAVI (mL/m<sup>2</sup>) | Baseline | 44.8 (34.7–56.7) | 61.6 (50.3–77.5) | 49.5 (40.1–69.9) | 76.0 (56.5–91.7) | <0.001 |
|  | 1-year | 37.7 (30.4–47.0) | 42.7 (34.0–56.4) | 58.8 (49.2–73.2) | 69.6 (54.0–89.2) | <0.001 |
|  | Δ (1yr–baseline) | -5.1 (-15.3–2.1) | -19.9 (-31.7–-6.6) | 7.8 (-1.4–17.2) | -0.1 (-16.1–10.0) | <0.001 |
|  | Within-group p† | <0.001 | <0.001 | 0.073 | 0.160 |  |
| E/e' | Baseline | 11.7 (9.1–14.6) | 20.7 (16.0–27.8) | 15.0 (10.8–16.5) | 24.0 (18.0–30.7) | <0.001 |
|  | 1-year | 10.0 (8.0–13.0) | 11.0 (8.8–13.9) | 19.6 (15.1–21.5) | 21.0 (16.0–29.4) | <0.001 |
|  | Δ (1yr–baseline) | -1.1 (-4.0–1.0) | -9.6 (-15.9–-5.0) | 5.2 (1.0–9.4) | -1.8 (-7.0–3.7) | <0.001 |
|  | Within-group p† | <0.001 | <0.001 | <0.001 | 0.010 |  |
| LASr (%) | Baseline | 17.6 (14.5–22.0) | 9.1 (7.0–12.0) | 16.1 (14.2–17.6) | 8.3 (6.2–10.8) | <0.001 |
|  | 1-year | 21.7 (17.4–26.4) | 19.2 (14.6–24.5) | 10.9 (9.3–13.8) | 8.8 (6.6–11.3) | <0.001 |
|  | Δ (1yr–baseline) | 3.0 (-0.9–7.6) | 9.2 (4.7–15.0) | -6.3 (-7.8–-1.9) | 0.8 (-2.1–3.0) | <0.001 |
|  | Within-group p† | <0.001 | <0.001 | <0.001 | 0.024 |  |
| LASI | Baseline | 0.7 (0.5–0.9) | 2.2 (1.6–3.3) | 0.8 (0.7–1.1) | 2.9 (1.9–4.4) | <0.001 |
|  | 1-year | 0.5 (0.3–0.7) | 0.6 (0.4–0.8) | 1.6 (1.4–2.0) | 2.3 (1.7–3.6) | <0.001 |
|  | Δ (1yr–baseline) | -0.2 (-0.4–0.0) | -1.5 (-2.7–-1.0) | 0.8 (0.7–1.0) | -0.3 (-1.6–0.7) | <0.001 |
|  | Within-group p† | <0.001 | <0.001 | <0.001 | <0.001 |  |
| PASP (mmHg) | Baseline | 28.0 (24.4–33.5) | 41.0 (30.0–51.0) | 30.0 (27.0–36.7) | 43.0 (33.0–53.4) | <0.001 |
|  | 1-year | 26.2 (22.6–31.2) | 28.0 (24.4–32.0) | 30.5 (26.6–36.3) | 37.0 (28.0–48.4) | <0.001 |
|  | Δ (1yr–baseline) | -1.8 (-6.2–3.4) | -9.0 (-19.2–-1.9) | 0.0 (-4.6–9.4) | -4.2 (-10.1–4.5) | <0.001 |
|  | Within-group p† | <0.001 | <0.001 | 0.398 | <0.001 |  |
| log NT-proBNP | Baseline | 6.4 (5.6–7.4) | 7.8 (7.0–8.6) | 7.4 (6.8–8.2) | 8.1 (7.3–8.9) | <0.001 |
|  | 1-year | 5.9 (4.9–6.7) | 6.3 (5.4–7.3) | 7.5 (6.7–7.8) | 7.7 (6.8–8.6) | <0.001 |
|  | Δ (1yr–baseline) | -0.3 (-1.1–0.0) | -1.2 (-2.5–-0.2) | 0.0 (-0.5–0.4) | -0.2 (-0.8–0.1) | <0.001 |
|  | Within-group p† | <0.001 | <0.001 | 0.831 | <0.001 |  |

Data are presented as median (interquartile range).
\* p value from Kruskal-Wallis test across all four groups; † Within-group p value from Wilcoxon signed-rank test (baseline vs. 1-year).
Abbreviations: E/e', ratio of early transmitral inflow velocity to early diastolic mitral annular velocity; LASI, left atrial stiffness index; LAVI, left atrial volume index; LASr, left atrial reservoir strain; LVEF, left ventricular ejection fraction; PASP, pulmonary artery systolic pressure.

### Clinical Outcomes

Over a median follow-up of 26.1 months (IQR, 14.9–40.1), 94 all-cause deaths, 51 cardiovascular deaths, and 97 HF hospitalizations occurred. Cumulative event rates differed significantly across the four groups for all three outcomes (log-rank p<0.001 for all; **Figure S2**). The cumulative incidences of all outcomes were highest in Group D, followed by Group C, and lowest in Groups A and B, which showed comparably low event rates. On multivariable analysis, Group D was independently associated with elevated risks of all-cause mortality (aHR 2.68, 95% CI 1.57–4.59), cardiovascular mortality (aHR 4.36, 95% CI 1.97–9.64), and HF hospitalization (aHR 3.83, 95% CI 2.22–6.60) (**Figure 3**). In Group B, the trend toward lower mortality was preserved, although its statistical significance was attenuated after multivariable adjustment. To quantify the incremental prognostic value of LASI trajectory group classification beyond its individual echocardiographic components, we compared C-statistics across multivariable Cox models incorporating interval changes in LASr (ΔLASr) and E/e’ (ΔE/e′), and LASI trajectory group classification sequentially (**Figure 4**). Addition of ΔLASr significantly improved discrimination for all three clinical outcomes, whereas ΔE/e′ alone provided only modest or non-significant incremental value. Incorporation of LASI trajectory group classification yielded the largest improvement in discrimination across all outcomes compared to the base model. Moreover, LASI trajectory group classification provided significant additive discrimination beyond models including either ΔE/e′ or ΔLASr (p<0.001 for all), indicating that the LASI trajectory classification captures prognostic information that is not fully explained by either component parameter alone.

**Figure 3.**
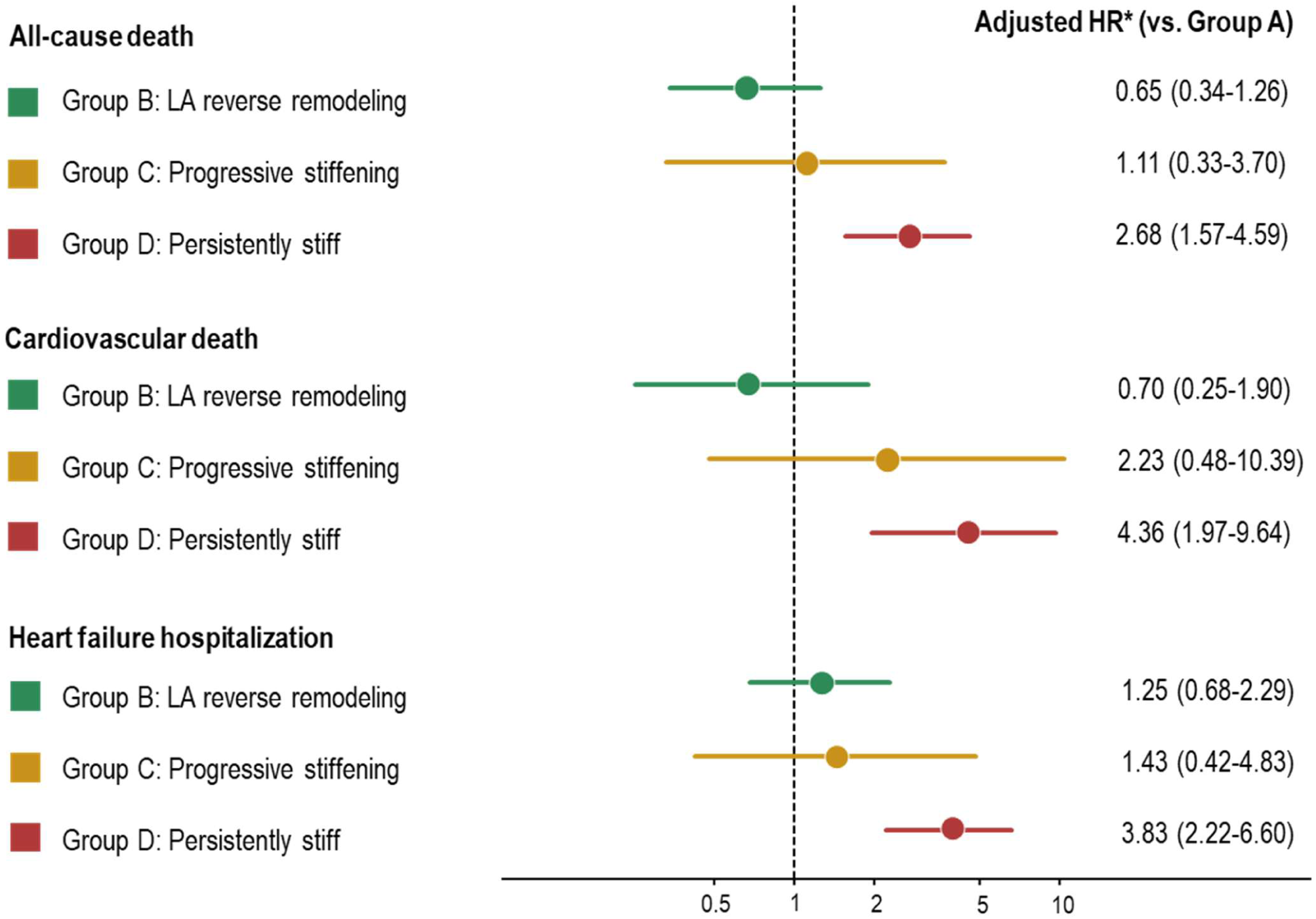
Hazard Ratios of Clinical Outcomes by LASI Patterns. Adjusted hazard ratios for all-cause death, cardiovascular death, and heart failure hospitalization according to LASI trajectory group, with Group A (persistently compliant) as the reference. Multivariable models were adjusted for age, sex, hypertension, diabetes mellitus, chronic kidney disease, atrial fibrillation, baseline LVEF, baseline LAVI, beta-blocker use, mineralocorticoid receptor antagonist use, and sodium-glucose cotransporter-2 inhibitor use. Data are presented as hazard ratio (95% confidence interval). HF, heart failure; LASI, left atrial stiffness index; LAVI, left atrial volume index; LVEF, left ventricular ejection fraction.

**Figure 4.**
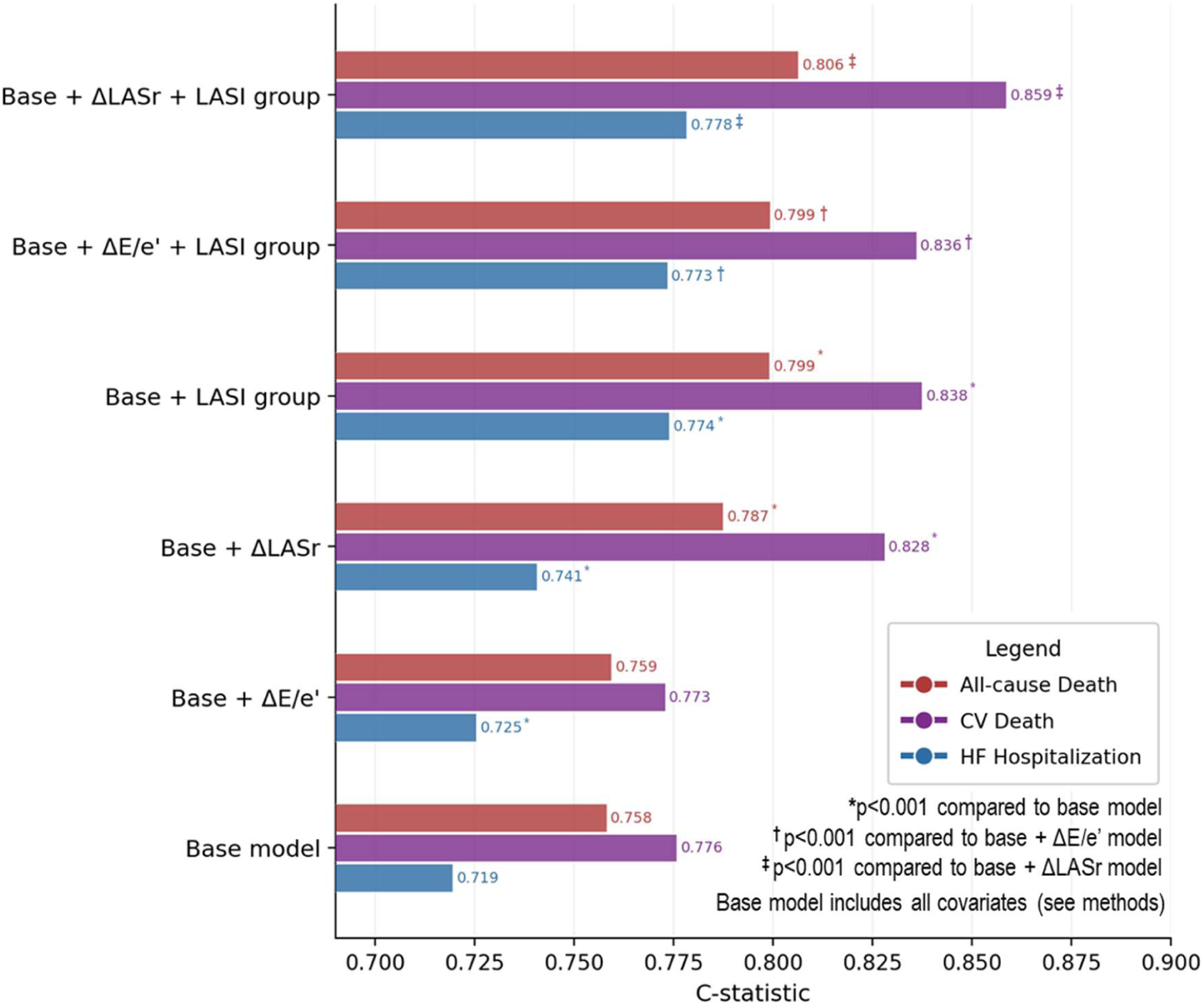
Incremental Prognostic Discrimination of LASI Patterns. C-statistics for all-cause death, cardiovascular death, and heart failure hospitalization across nested multivariable Cox models. The base model included all covariates listed in the Methods. The serial change in E/e′ (ΔE/e′), the serial change in LA reservoir strain (ΔLASr), and the LASI trajectory group classification were added sequentially to the base model. *p<0.001 compared with the base model; ^†^p<0.001 compared with the base + ΔE/e′ model; ^‡^p<0.001 compared with the base + ΔLASr model. LASI, left atrial stiffness index; LASr, left atrial reservoir strain.

The prognostic significance of LASI was further examined across LVEF recovery strata (**Figure 5**). When patients were cross-classified by LV recovery status (ΔLVEF ≥10% vs. <10%) and one-year LASI status, high LASI at one year was consistently associated with higher event rates regardless of whether LV recovery was achieved (**Figure 5A**). Similarly, when stratified by one-year LVEF category (<40%, ≥40 to <50%, and ≥50%), high LASI at one year was associated with higher event rates within each LVEF stratum (**Figure 5B**). Notably, even among patients with fully recovered LV function (one-year LVEF ≥50%), persistent LASI elevation conferred a markedly elevated risk of all-cause mortality (aHR 10.82, 95% CI 2.61–44.82).

**Figure 5.**
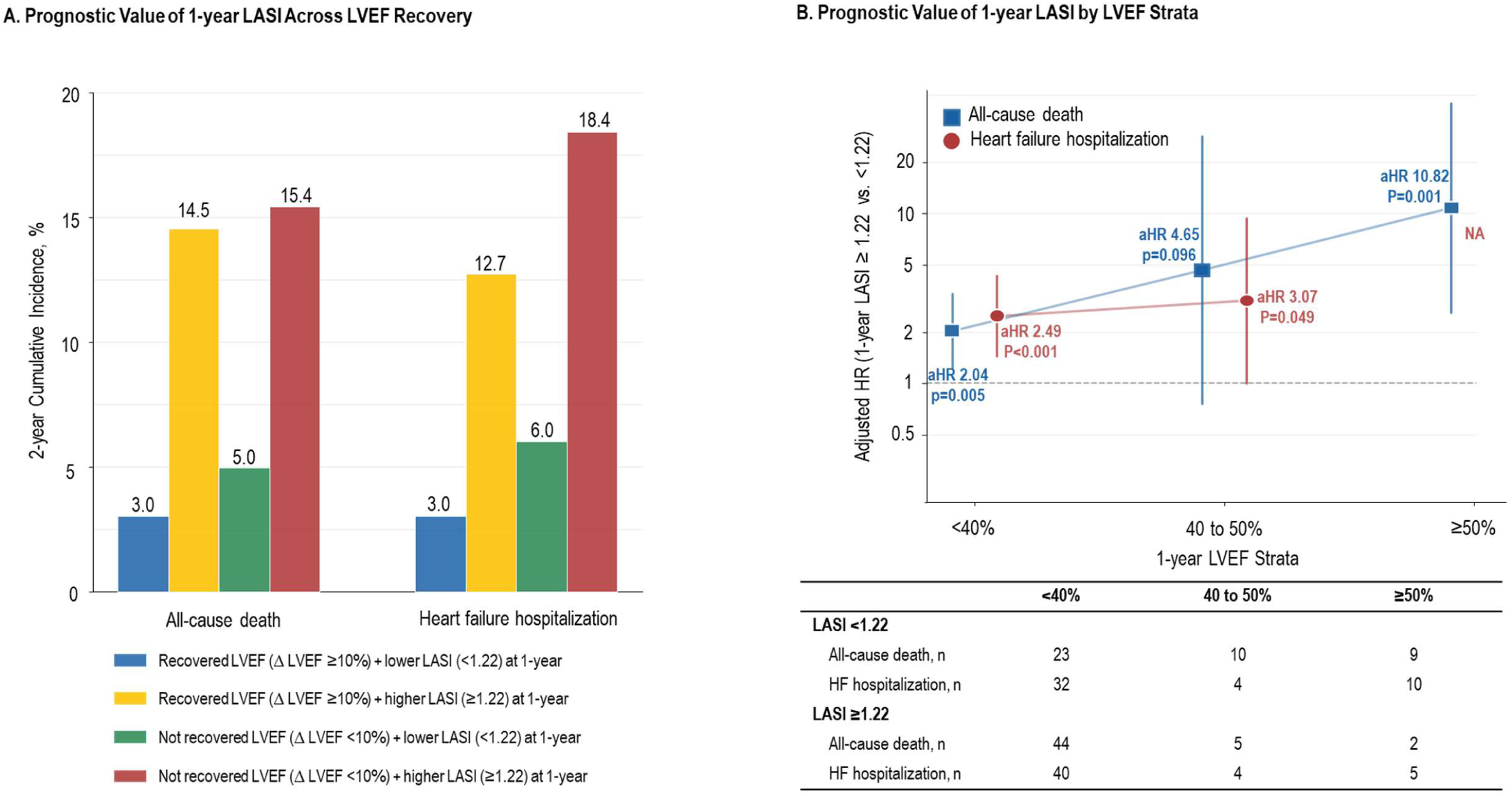
Prognostic Value of LASI pattern Across LVEF Recovery. (A) Two-year cumulative incidence of all-cause mortality and HF hospitalization, cross-classified by LV recovery status (ΔLVEF ≥10% vs. <10%) and one-year LASI status (below vs. at or above the cohort median). (B) Event counts for all-cause mortality and HF hospitalization, stratified by one-year LVEF category (<40%, ≥40 to <50%, and ≥50%) and one-year LASI status. In the LVEF ≥50% stratum, the hazard ratio for HF hospitalization could not be reliably estimated due to the limited number of events. HF, heart failure; LASI, left atrial stiffness index; LVEF, left ventricular ejection fraction.

### Risk of New-onset Atrial Fibrillation by LASI Trajectory

Among 786 patients in sinus rhythm without a history of AF at baseline, 39 new-onset AF events occurred. Cumulative incidence of new-onset AF differed significantly across the four LASI trajectory groups (log-rank p<0.001; **Figure 6A**). Group A showed the lowest event rate, whereas Groups C and D exhibited the highest rates. On a multivariable Cox regression model with Group A as the reference, all three remaining groups were independently associated with elevated risk of new-onset AF. The risk was most pronounced in Group C (aHR 6.84, 95% CI 2.03–23.08), the group characterized by progressive stiffening of an initially compliant LA, followed by Group D (aHR 5.10, 95% CI 2.12–12.24) and Group B (aHR 2.95, 95% CI 1.21–7.20).

**Figure 6.**
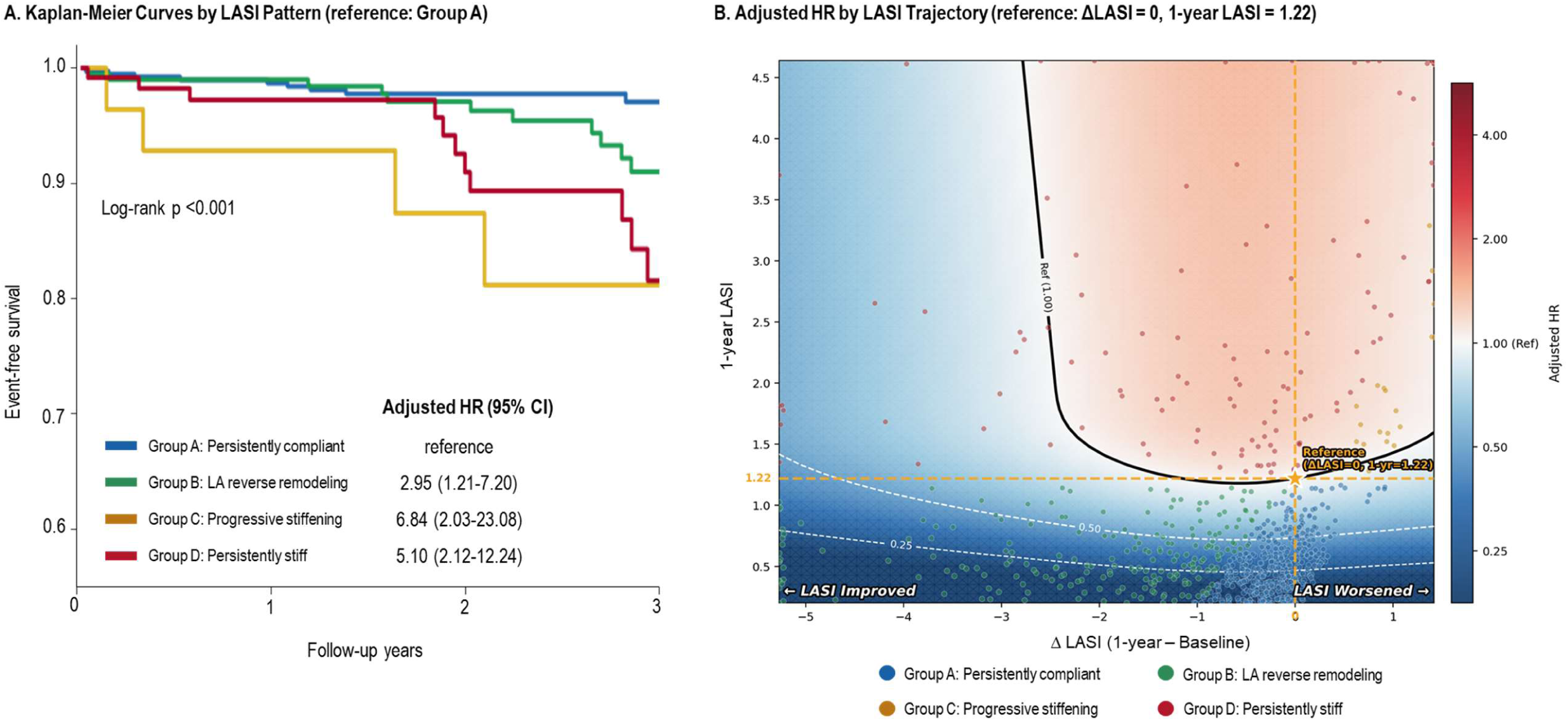
Risk of New-onset Atrial Fibrillation According to LASI Pattern. (A) Kaplan-Meier curves and adjusted hazard ratios for new-onset AF according to LASI trajectory group, among the patients in sinus rhythm without a history of AF at baseline (n=786). Group A (persistently compliant) served as the reference. (B) Adjusted hazard ratio surface for new-onset AF as a function of one-year LASI (y-axis) and ΔLASI (x-axis), modeled using a multivariable Cox regression model with restricted cubic splines for both variables. The reference point was set at no change in LASI (ΔLASI = 0) and the cohort median of baseline LASI. Multivariable adjustment was identical to that used in the primary analysis. AF, atrial fibrillation; LASI, left atrial stiffness index.

The joint association of ΔLASI and one-year LASI with new-onset AF was further evaluated using restricted cubic splines for both variables, with hazard ratios visualized as a heatmap (**Figure 6B**). Higher one-year LASI and worsening ΔLASI were both associated with increased AF risk in a graded manner. Group D was distributed across the upper region of the plot corresponding to elevated one-year LASI, and Group C clustered in the upper-right corner reflecting both LASI worsening and elevated one-year LASI, where the highest hazard ratios were observed. Conversely, lower one-year LASI accompanied by improvement in LASI was associated with reduced AF risk. Group B was distributed in the lower-left region of the heatmap, where cases with pronounced LASI improvement demonstrated AF risk comparable to that of Group A.

## DISCUSSION

In this multicenter real-world registry of patients with HFrEF treated with ARNI-based GDMT, we characterized four distinct LASI trajectory patterns based on concurrent baseline and one-year echocardiographic assessments. The principal findings are as follows. First, among the four LASI trajectory groups, persistently stiff LA group (Group D) was independently associated with elevated risks of all-cause mortality, cardiovascular mortality, and HF hospitalization, whereas patients with LA reverse remodeling (Group B) showed risks comparable to the persistently compliant group (Group A). Second, the prognostic value of one-year LASI persisted even among patients with LV functional recovery, indicating that LA stiffness captures residual hemodynamic burden not reflected by LVEF recovery alone. Third, in the sinus rhythm subset, the progressive stiffening group (Group C) showed an increased risk of new-onset AF, and bivariate modeling of one-year LASI and ΔLASI demonstrated that pronounced LASI improvement was associated with an attenuated AF risk, comparable to that of patients with persistently compliant LA, who had the lowest AF risk. Together, these findings suggest that serial LASI assessment provides prognostic information beyond LV-centric metrics in HFrEF and identifies clinically relevant phenotypes for risk stratification. To our knowledge, this is the first study to evaluate baseline-to-one-year LASI trajectories and to demonstrate their prognostic implications in patients with HFrEF.

LV reverse remodeling, particularly LVEF improvement, remains the most important indicator of treatment response in HFrEF. Patients with LVEF recovery generally have substantially better outcomes than those with persistently reduced LVEF; however, they may continue to experience HF events, abnormal biomarker profiles, and adverse outcomes, suggesting that LV recovery is an important but incomplete marker of global disease modification.^4,5,18^ Accordingly, indices reflecting residual hemodynamic burden, diastolic stress, and atrial remodeling may complement LV-centric measures and improve risk stratification after treatment.^18^ In the present study, elevated one-year LASI was independently associated with worse outcomes across all LVEF recovery strata, including patients who achieved LVEF normalization, directly illustrating the incremental value of LA stiffness assessment beyond LV function.

LASI provides a physiologically integrated approach for assessing LA hemodynamic burden. Conventional diastolic parameters, such as E/e′ and LAVI, each reflect only one dimension of the LA–LV interaction: E/e′ estimates LV filling pressure, whereas LAVI represents cumulative structural remodeling but may be insensitive to dynamic changes in LA mechanical function.^19^ In contrast, LASI combines E/e′ with LASr, a marker of LA reservoir compliance, simultaneously capturing both the pressure load imposed on the LA and the mechanical response of the atrial wall, which may explain its established prognostic value in HF populations.^11,12^ The 2025 ASE guidelines for diastolic function assessment formally incorporated LASr as a recommended parameter for estimating LV filling pressure and acknowledged the potential usefulness of LASI, underscoring the growing clinical recognition of LA strain-based indices beyond conventional volumetric and Doppler parameters.^17^

Importantly, the present study demonstrates that LASI trajectory group classification provides greater incremental prognostic discrimination than either of its individual echocardiographic components. Furthermore, LASI trajectory group classification provided significant additive discrimination beyond models incorporating either ΔE/e′ or ΔLASr, indicating that the four-category trajectory classification captures prognostic information not fully explained by either component parameter alone. These findings extend recent work by Abou Kamar et al., who demonstrated that although repeatedly measured LASr was independently associated with adverse events in HFrEF, temporal trajectories of LASr did not provide incremental prognostic value over single baseline measurements.^20^ The superior performance of LASI trajectory classification in the present study likely reflects the composite nature of the index by simultaneously integrating changes in both LV filling pressure (E/e′) and LA reservoir function (LASr). As a result, LASI trajectory may capture pathophysiological patterns that neither component alone can fully characterize.

New-onset AF is a clinically important complication in HF that worsens symptoms, impairs cardiac output, increases thromboembolic risk, and contributes to HF progression. Elevated LA filling pressure, impaired reservoir function, atrial stretch, and structural remodeling collectively provide the substrate for AF development. Prior studies have demonstrated that LA strain-based parameters independently predict new-onset AF in patients with HF and sinus rhythm, supporting the importance of LA functional assessment beyond LA size alone.^7,8,21^ However, these studies have mainly evaluated single time-point measurements, and whether serial changes in LA stiffness are associated with subsequent AF risk has not been established. To our knowledge, the present study is the first to demonstrate that baseline-to-follow-up changes in LASI are independently associated with new-onset AF risk in patients with HFrEF, adding a temporal dimension to the known relationship between LA stiffness and AF susceptibility. Among the trajectory groups, progressive stiffening of an initially compliant LA (Group C) was associated with the marked increase in new-onset AF risk, comparable to even that of patients with persistently stiff LA (Group D). Persistently stiff LA likely represents a chronic and relatively advanced maladaptive state in which structural and fibrotic remodeling has reached a plateau, leaving fewer acutely reversible substrates for incident AF. In contrast, progressive stiffening from an initially compliant state may reflect a dynamic transition phase, during which ongoing increases in LA pressure, emerging interstitial fibrosis, and accelerating electrical remodeling collectively create an acutely arrhythmogenic substrate. This interpretation suggests that monitoring the direction of LASI change, rather than its absolute value at a single time point, may improve identification of patients at heightened arrhythmic risk. However, because the Group C cohort was relatively small, this observation requires further prospective validation.

From a clinical perspective, the current study findings suggest that serial LASI assessment may refine follow-up strategies in patients with HFrEF receiving ARNI-based therapy. While LVEF improvement remains the primary treatment target, persistent or worsening LA stiffness may identify patients with residual hemodynamic burden who require closer clinical surveillance for HF events and rhythm disturbances, even after apparent LV recovery. As the use of LASr expands from research settings into real-world clinical practice, incorporation of LASI into standard follow-up assessment may become increasingly feasible and clinically relevant. Whether serial LASI-guided management strategies can improve clinical outcomes warrants investigation in prospective trials.

Several limitations should be acknowledged. First, this was a retrospective observational registry analysis, and residual confounding cannot be excluded despite multivariable adjustment. Second, a considerable proportion of initially eligible patients were excluded due to the absence of serial echocardiographic data; thus, the prognostic implications of LASI trajectory may differ in patients without serial assessments. Third, because validated cutoffs for serial LASI in ARNI-treated HFrEF have not been established, the cohort median was used as a dichotomization threshold; this threshold should be regarded as a research-based stratification rather than a universal clinical cutoff. Finally, external validation in independent cohorts is required to confirm the generalizability and clinical utility of serial LASI trajectory classification for risk stratification in HFrEF.

## CONCLUSION

In patients with HFrEF receiving ARNI-based therapy, four distinct LASI trajectory patterns were identified, each associated with a differential prognosis. Persistently stiff LA was independently associated with a substantially increased risk of mortality and HF hospitalization, whereas LA reverse remodeling conferred a prognosis comparable to that of persistently compliant LA. The prognostic significance of LASI trajectory persisted across LVEF recovery status and provided incremental prognostic values over conventional indices. Notably, progressive LA stiffening was associated with an increased risk of new-onset AF. These findings support the role of serial LASI assessment as an integrative prognostic tool for risk stratification in HFrEF beyond LV-centric metrics.

## PERSPECTIVES

## COMPETENCY IN PATIENT CARE AND PROCEDURAL SKILLS

In patients with HFrEF treated with ARNI-based therapy, serial assessment of the left atrial stiffness index identifies distinct prognostic phenotypes and detects residual hemodynamic risk that persists despite improvement in left ventricular ejection fraction.

## TRANSLATIONAL OUTLOOK

Prospective studies are needed to determine whether longitudinal monitoring of left atrial stiffness can guide intensification of therapy and improve outcomes in patients with HFrEF and persistent left atrial dysfunction.

## Data Availability

The data underlying this study cannot be made publicly available due to ethical restrictions set by the IRB of the study institution; i.e., public availability would compromise patient confidentiality and participant privacy. Please contact the corresponding author to request the minimal anonymized dataset.

